# Magnitude of work-related musculoskeletal disorders and ergonomic risk practices among medical laboratory professionals in Northwest Ethiopia: A cross-sectional study

**DOI:** 10.1101/2023.01.09.23284343

**Authors:** Mekuriaw Temeche Alibsew, Habtamu Molla, Melashu Balew Shiferaw, Abay Sisay Misganaw

## Abstract

**Objective:** This study aimed to assess the magnitude of work-related musculoskeletal disorders (WMSDs) and ergonomic risk practices among medical laboratory professionals in North West Ethiopia.

**Design:** Facility-based cross-sectional study

**Methods:** Cross-sectional study design was employed among medical laboratory professionals (MLPs). The Nordic musculoskeletal questionnaire was adopted and used. In addition to questionnaires about socio-demographic characteristics and ergonomic risk practice, one-to-one interviews, and a direct observational checklist were used. Data was entered into Epi Data 3.1 and then exported and analyzed using SPSS version 25.0. Logistic regression analysis was used to estimate the 95% CI (AOR) at a cut-off value of p <0.05 for statistically significant tests.

**Results:** A total of 238 MLPs participated in the study. The magnitude of WMSDs was 116(48.7%). The most affected body parts were the lower back (20.6%) and wrists (16.4%). The magnitude of WMSDs among government-owned hospitals was the highest (56.4%). 67.6% MLPs never heard about ergonomics. The general mean score of workstations was 2.28. Ergonomic risk practices like repetitive movement and doing of high workload were significantly associated with WMSDs.

**Conclusion:** The current findings revealed a high magnitude of WMSDs that strongly need applying preventive action before body symptoms developed. Improving and renovating workplace design and enhancing awareness of MLPs were the necessary measures to control ergonomic risk factors.

## Introduction

Musculoskeletal disorders (MSDs) are injuries of the muscles, nerves, tendons, joints, cartilage, and spinal discs.^1^ The main risk factors for MSDs development among MLPs are ill-structured jobs, poor posture at work, poor workstation design, prolonged working time, and repetitive movements. ^2^ Differently, routine laboratory examination needs procedures such as pipetting, working at microscopes, and operating microtomes that requires repetitive motion, which is risky. ^3^

A work setting causes workplace risk factors for MSDs such as shoulder and backache, joint pain and muscle fatigue, this suffering mainly lies on health care professionals comprising MLPs.^4^ Particularly, medical laboratory technicians have high a magnitude of WMSDs and the major affected areas were neck, shoulder, elbow, and hand. ^5^ Hence, the magnitude of WMSDs among MLPs is high which ranging from 40 to 60%.^6^

One of the pertinent causes for increased risk of WMSDs development among MLPs is the lack of application of ergonomic principle.^2^ Prolonged exposure to ergonomic risk factors can also cause damage to a worker’s body and lead to WMSDs.^7^ Nevertheless, there are limited studies about ergonomic hazards that ultimately lead to musculoskeletal problems among MLPs in Ethiopia.

This study aimed to assess the magnitude of work-related musculoskeletal disorders (WMSDs) and ergonomic risk practice among MLPs working in Bahirdar, Northwest Ethiopia from February 1, 2021 to June 30, 2021. We hypothesized that there is no significant association between the occurrence of WMSDs and ergonomic risk practice among MLPs in Northwest Ethiopia.

## Methods

### Study design and Setting

Cross-sectional study was conducted to assess the magnitude of WMSDs and ergonomic risk practice among medical laboratory professionals of Bahirdar, Northwest Ethiopia by incorporating the quantitative and direct observational techniques were used from February 1, 2020 to June 30, 2021. Bahirdar city had three government owned hospitals and four private owned hospitals in addition ten government owned health centres. Ten private owned clinics also included in the study. Furthermore, the city had Amhara public health institute (APHI) accredited by the Ethiopian National Accreditation Office (ENAO) in all laboratory tests since 2018G.C in accordance with the ISO 15189:2012 standard. ^8^ According to the health and health indicators of Amhara regional state health bureau, the number of MLPs in the city is 264.

### Sample size and sampling techniques

All MLPs who were working in health facilities of Bahirdar that fulfils the inclusion criteria were taken as study subject. Thus, no need of sample size calculation for the determination of the study variables. Accordingly, two hundred thirty eight (238) MLPs working in health facilities of Bahirdar city were included in the study. Totally 27 health facilities medical laboratory departments were assessed. One hundred fifty six workstations that laboratory staffs were on work during data collection moment were selected purposively. The health facilities were selected using purposively by taking all the health facilities having laboratory service during the study period. Census sampling technique employed for those MLPs who actively engaged in the laboratory diagnosis of government and private owned health facilities as well as and who had fixed working hours.

### Data collection

The questionnaires, checklists and interviews, data collection tools were interrelated to assess the magnitude about association of WMSDs and ergonomic risk practices among MLPs. The questionnaires were consists of socio-demographic characteristics, and pain and discomfort. Nordic Musculoskeletal questionnaire which is Nordic measurement that divides human body into nine body regions, which may affect by WMSDs, institutional factors, and workplace factors. ^9, 10^ Direct observational checklist was used to determine the condition of standing bench, sitting bench, computer, pipetting, microscope, and chair and micromanipulation workstations. Moreover, tools like Paper, pencil, a folding rule, and a camera was used for workplace observations and for identification of ergonomic hazards using PLIBEL checklist. ^10^ One to one interviews using semi-structured questions with all medical laboratory managers were carried out. This data collection tools were pretested in Debretabor comprehensive specialized hospital.

### Data Quality Assurance

The data quality was assured before, during and after the data collection. Before data collection, the questionnaires were pre tested in Debretabor referral hospital to evaluate its clearness and applicability according to the objective of the study. After the data collection, the collected data was rechecked for its completeness and consistency by the supervisor and principal investigator. The completeness of the questionnaire and its clarity was mainly rechecking by principal investigator. Nordic musculoskeletal questionnaire had an anatomical diagram of nine body regions like neck, shoulder, upper and lower back, hands/wrists, arms, knee, thighs and feet to make easy for study participants on correctly identifying the presence of musculoskeletal symptoms.

### Patient and public involvement

Neither patients nor the public were involved in the design of this study.

### Data analysis and interpretation

The data was edited, coded, and double entered into SPSS (version 25, SPSS Inc., Chicago, IL, USA) for cleaning and analysis. Odds ratio with 95% confidence interval was used to measure the association between the independent variables with the dependent variables. Results were summarized in texts, frequency tables and graphs. The association between report of WMSDs and respondents’ socio demographic and ergonomics risk practice data were checked by using the binary logistic regression analysis. In addition, chi-square assumption was checked. For statistically significant tests and to count in the last logistic model the cut off value was P < 0.25. In the two variables (bivariate) analysis, the crude OR (COR) of reports of WMSDs was estimated by covariates; all covariates were categorical. Furthermore, to clear confounders and to check for interactions, multiple logistic regression analysis was used to estimate the adjusted OR at a cut off value of p <0.05 for statistically significant using Hosmer and Lemeshow goodness of fit test.

### Medical laboratories Workstations

Standard ruler was used to measure the selected workstation. The measured values (centimetres) were converted into suitable units (meter). At the selected workstation, direct observational data was recorded while medical laboratory personnels were at work. Each workstation was evaluated using assessment checklist. ^10^ The collected data were summarized in texts and frequency tables.

If the workstations had the item when assessed, then it was score “1” (representing “yes”). While, if the workstation had not the required item when assessed, then it was score (“0” representing “no”). Suboptimal conditions and risk of injury represented by “no” answers. The sum of all item score gives total score of workstation. The sum of all item scores was dividing the total score of workstation. The total score of each workstation was divided by the number of items checked; the final score was expressed as a percentage. ^11^

### Categorizing of medical laboratory workstations

Workstations of medical laboratories was categorized according to the following components: the standard height of standing bench is 0.91m, the standard height of sitting bench is 0.76m, laboratory chairs, microscopes (in bacteriology, haematology, parasitology, and urine analysis), pipettes, micromanipulation (of vials, forceps, cap openers, and tube holders), miscellaneous (i.e., platforms, storage space, storage closets, bins, and racks), and computer workstation. ^11, 12^

### Oprational Definitions

Work load: - the number of patients served in a day by one professional is greater than 35, it considered as loaded. ^13^.

Work related musculoskeletal disorders: - conditions when the immediate working environment intensifies injuries of muscles, nerves, tendons, joints, cartilage, and spinal discs for at least 2 to 3 days during the last 12 months on any body parts. These symptoms often come along at the work and go away at the time of rest and symptoms continue after work ends. These symptoms often appear at work that disappears during rest and symptom continues after work ends. ^1, 14, 15^

## Results

### Socio-demographic characteristics of the study participants

Out of the 264 questionnaires distributed to health facilities of Bahirdar, 238(90%) complete questionnaires returned and the majority of them,139(58.4%) were from government owned health facility. The age of MLPs ranging from 20 to 58 years and the mean age was 30.81years with standard deviation = 5.704 years additionally, 197(82.8%) participants were in an age range of 20 to 35 years. The mean work experience was 7.77(SD=4.982) and 153(64.3%) professionals had a work experience of greater than 5 years in their medical laboratory work place (Table 1).

**Table 1.** Demographics of MLPs working in Bahirdar, Ethiopia, 2021(n=238)

| Variables | Category | Frequency | % |
| --- | --- | --- | --- |
| Status of health facilities | Public hospital | 139 | 58.4 |
|  | Public health center | 30 | 12.6 |
|  | Private hospital | 38 | 16.0 |
|  | Private clinic | 31 | 13.0 |
| Sex | Male | 147 | 59.7 |
|  | Female | 91 | 38.2 |
| Age in years | 20-35 | 197 | 82.8 |
|  | 36-60 | 41 | 17.2 |
| Marital status | Single | 135 | 56.7 |
|  | Married | 97 | 40.8 |
|  | Divorced | 6 | 2.5 |
| Educational status | College diploma | 96 | 40.3 |
|  | First degree | 116 | 48.7 |
|  | Second degree | 26 | 10.9 |
| Work experience | <5 years | 86 | 35.7 |
|  | 5-15 years | 137 | 57.6 |
|  | >15 years | 16 | 6.7 |
| Monthly income in Ethiopian birr | <4501 | 58 | 24.4 |
|  | 4501-7501 | 127 | 53.4 |
|  | >7501 | 53 | 22.3 |

### Magnitude distribution of WMSDs among body parts of MLPs

Over the past 12 months, about half 116(48.7%) of clinical laboratory staffs reported WMSDs at least one of the nine body parts, they reported 274 WMSDs and the most affected body parts were the lower back 49 (20.6%) (Figure 1).

**Figure 1.**
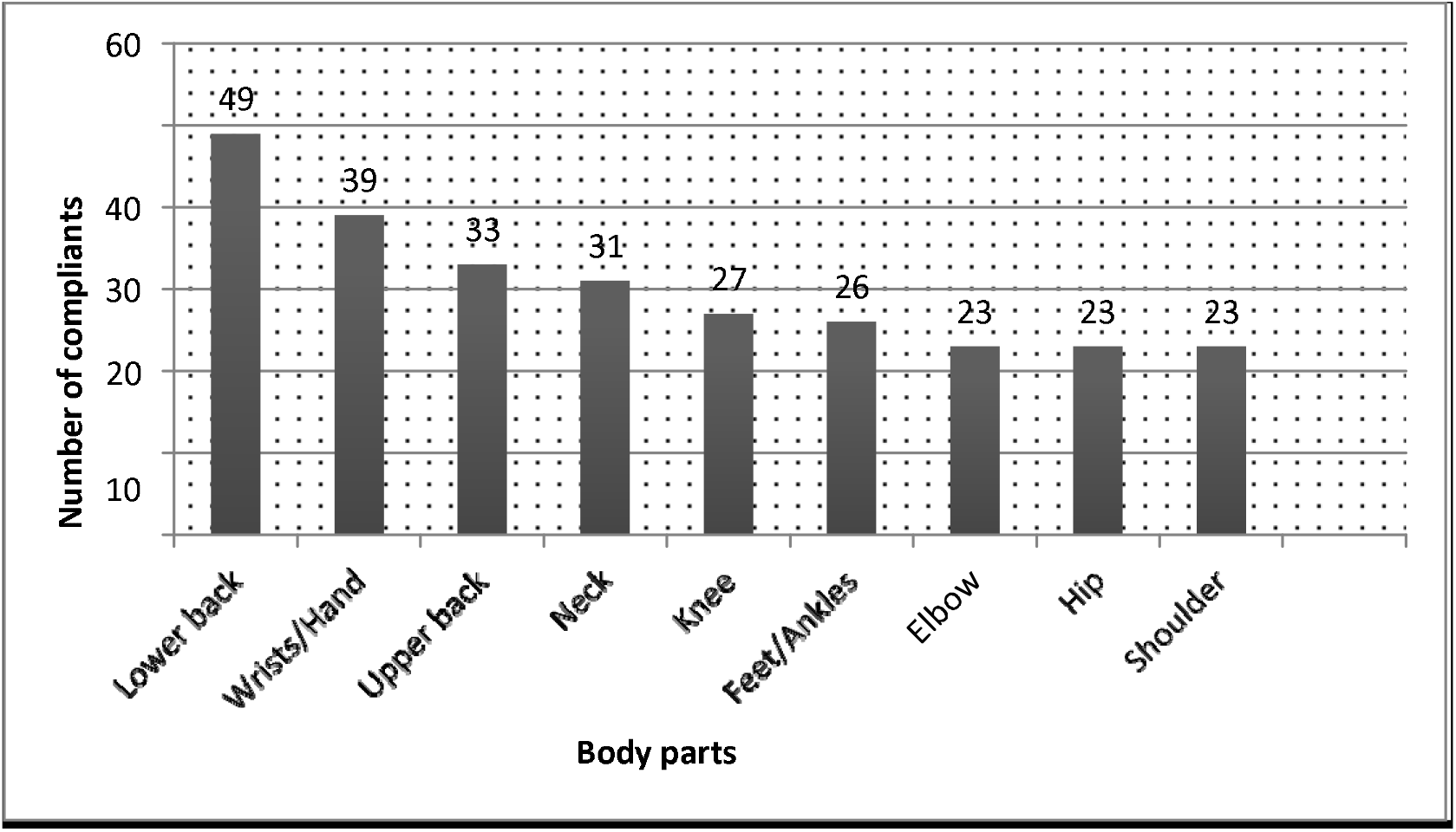
Number of body complaints among MLPs working in Bahirdar, Ethiopia, 2021

### Frequency of pain and discomfort within medical laboratory professionals

One hundred one (42.4%) of participants took medication because of WMSDs. The most affected was the wrist 18(7.6%) and the ankles at 16 (6.7%). The cause of the disorder that was judged by the respondents were repetitive work 83(34.9%) and awkward posture 26(10.9%). Of those who had disorders, 83(38.4%) suffers pain 2 to 3 times per week. Thirty-nine (16.4%) got medication because of the disorders. Among those 25(10.5%) took 265 sick leave days that range from 2 to 90 days with mean of 10.6 days (SD= 17.543) (Table 2).

**Table 2.** Pain and discomfort among MLPs working in Bahirdar, Ethiopia, 2021

| Variables | Category | Frequency(n=238) | (%) |
| --- | --- | --- | --- |
| WMSDs within a year | Yes | 116 | 48.7 |
|  | No | 122 | 51.3 |
| Likely cause of the pain | Repetition | 82 | 34.5 |
|  | Awkward posture | 26 | 10.9 |
|  | Vibration | 5 | 2.1 |
|  | Static loading | 1 | 0.4 |
|  | Forceful exertion | 2 | 0.8 |
| Medication for pain | Yes | 39 | 16.6 |
|  | No | 77 | 32.3 |
| Frequency pain of WMSDs per week | 1-2 | 75 | 31.5 |
|  | >2 | 34 | 14.3 |
| sick leave | Yes | 25 | 10.5 |
|  | No | 91 | 38.1 |

### Magnitude distribution of WMSDs and types of health facility

MLPs who are working in government-owned hospitals report the highest annual magnitude of WMSDs 62(56.9%) followed by private hospitals 18(15.5%). Low back pain was highest among MLPs who work in public hospitals 28(57.2%) followed by private hospitals 8(16.3%) (Table 3).

**Table 3.** Musculoskeletal complaints of MLPs working in Bahirdar, Ethiopia, 2021

| Type of Musculoskeletal complaint | Ownership of health facilities |  |  |  |  |
| --- | --- | --- | --- | --- | --- |
|  | Gov't owned hospital | Gov't owned HC | Private hospital | Private clinic | Others |
|  | N (%) | N (%) | N (%) | N (%) | N (%) |
| WMSDs | 62(56.9) | 16(17.8) | 18(15.5) | 15(12.9) | 5(4.2) |
| Neck pain | 15(4.8) | 5(16.1) | 5(16.1) | 4(3.4) | 2(1.7) |
| Shoulder pain | 12(51.2) | 5(2.2) | 2(8.7) | 3(13) | 1(4.3) |
| Elbow/forearm pain | 14 (60.9) | 4(17.4) | 2(8.7) | 3(13) | 0(0.0) |
| Wrist/hand pain | 18(46.2) | 6(15.4) | 5(12.8) | 9(23.1) | 1(2.6) |
| Upper back pain | 19(59.4) | 7(21.9) | 2(6.3) | 3(9.4) | 2(6.3) |
| Lower back pain | 28(57.2) | 7(14.3) | 8(16.3) | 5(10.2) | 1(2.0) |
| Hip/thigh pain | 10(43.5) | 8(34.8) | 3(13.0) | 1(4.3) | 1(4.3) |
| Knees/leg pain | 11(40.7) | 4(14.8) | 4(14.8) | 6(22.2) | 2(8.7) |
| Ankle pain | 11(42.3) | 6(23.0) | 1(3.8) | 6(23.0) | 2(7.7) |

### Behavioural and individual factors of MLPs

#### Individual Characteristics of the medical laboratory professionals

Most professionals 221(92.9%) had normal BMI (18.5-24.9kg/m^2^) and the mean BMI was a mean of 1.94 (SD=0.260). 153(64.3%) professional staffs have no a habit of doing physical exercise whereas 85(35.7%) had an experience. Of those 51(21.4%) doing physical exercise two times per week. 15(6.3%), 8(3.4%), 12(5.0%) laboratory staffs had medical history of systemic illness, Symptoms related to WMSDs before engaged in this job, and were diagnosed for bone disease respectively(Table 4).

**Table 4.**
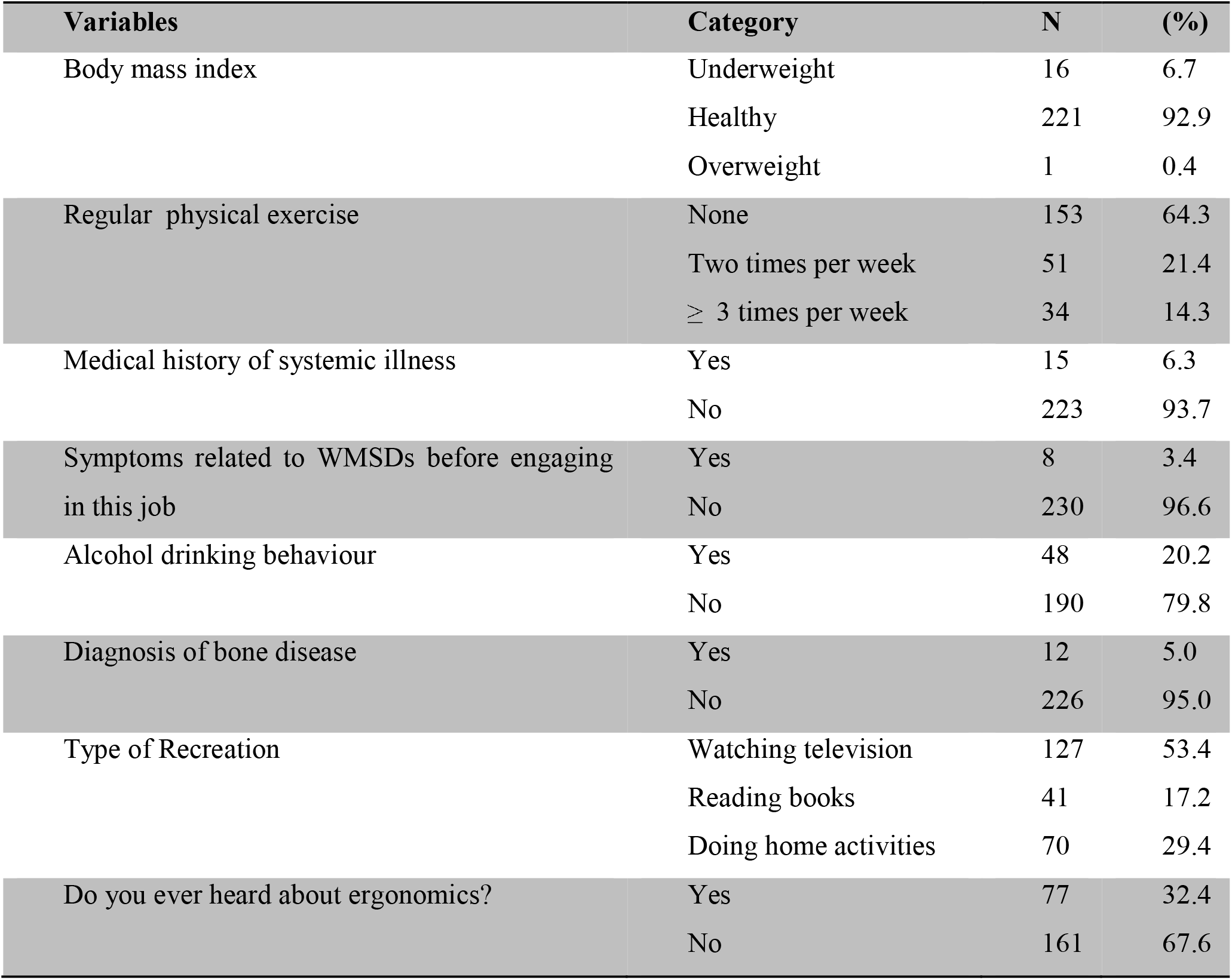
Individual characteristics of MLPs in Bahirdar, Ethiopia, 2021

### Ergonomic risk practices

The mean of weekly working days were 5.72(SD=1.019) and the mean of weekly working hours were 47.64(SD=10.42). Specifically, more than half of Medical laboratory staff 129(54.2%), and 140(58.8%) worked for more than five days and 41hours per week respectively. Furthermore, there were no working breaks other than lunchtime on regular laboratory work for one hundred forty (58.8%) medical laboratory individuals. Among the rest 66(27.7%) took a break of less than 15 minutes. One hundred thirty eight (58.0%) and 120(50.4%) respondents did highly loaded work sometimes and repetitive movements within <30 seconds respectively. One hundred thirty two (55.5%) of MLPs had overtime work. The study showed almost all 230(96.6%) of MLPs were permanently employed. 123(51.7%) respondents were work on public hospitals. Among MLPs 55(23.1%) were assigned on Parasitology and urinalysis section, 41(17.2%) Hematology, 36(15.1%) Clinical chemistry, and 62(26.1%) had miscellaneous (Table 5).

**Table 5.** Workplace characteristics of MLPs working in HFs of Bahirdar, Ethiopia, 2021

| **Variables** | **Category** | **Frequency** | **(%)** |
| --- | --- | --- | --- |
| working day in the week | ≤5 | 109 | 45.8 |
|  | >5 | 129 | 54.2 |
| working hours in the week | <40 | 98 | 41.2 |
|  | ≥40 | 140 | 58.8 |
| Presence of working breaks excluding lunchtime | No | 140 | 58.8 |
|  | ≤ 15 minutes | 66 | 27.7 |
|  | >15 minutes | 32 | 13.4 |
| Doing high-loaded work | never | 14 | 5.9 |
|  | sometimes | 138 | 58.0 |
|  | always | 86 | 36.1 |
| Repetitive movement within < 30 seconds | never | 31 | 13.0 |
|  | sometimes | 120 | 50.4 |
|  | always | 87 | 36.6 |
| Number of hours in overtime work per week | No | 106 | 44.5 |
|  | 1-15 | 82 | 34.5 |
|  | 16-30 | 41 | 17.2 |
|  | 31-45 | 9 | 3.8 |
| Rest between overtime work | No | 76 | 31.9 |
|  | 4-16 | 36 | 15.1 |
|  | 17-28 | 12 | 5.0 |
|  | 29-40 | 5 | 2.3 |
|  | >40 | 3 | 1.3 |
| Hours spent in second job per day | No | 172 | 72.2 |
|  | 1-5 | 45 | 18.9 |
|  | 6-10 | 18 | 7.6 |
|  | >10 | 3 | 1.3 |
| Number of hours in housework | No | 50 | 21.0 |
|  | 1-4 | 118 | 49.8 |
|  | 5-8 | 65 | 27.3 |
|  | 9-12 | 5 | 2.1 |
| Number of hours carrying children <5 | No | 126 | 52.9 |
|  | 1-4 | 76 | 31.6 |
|  | 5-8 | 33 | 13.9 |
|  | 9-12 | 3 | 1.3 |
| Employment status | Contract | 8 | 3.4 |
|  | Permanent | 230 | 96.6 |
| Status of health facility | Public hospital | 123 | 51.7 |
|  | Public health center | 30 | 12.6 |
|  | Private hospital | 38 | 16.0 |
|  | Private clinic | 32 | 13.4 |
|  | Others | 15 | 6.3 |
| Section of laboratory dept. | Clinical chemistry | 36 | 15.1 |
|  | Hematology | 41 | 17.2 |
|  | Bacteriology | 20 | 8.4 |
|  | Parasitology and urinalysis | 55 | 23.1 |
|  | ART laboratory | 8 | 3.4 |
|  | Blood bank | 16 | 6.7 |
|  | Miscellaneous | 62 | 26.1 |
| Workstations of MLPs | Standing bench | 34 | 14.3 |
|  | Seating bench | 42 | 17.6 |
|  | Laboratory chairs | 28 | 11.8 |
|  | Microscope | 29 | 12.2 |
|  | Pipettes | 28 | 11.8 |
|  | Micromanipulation | 11 | 4.6 |
|  | Computer | 53 | 22.3 |
|  | Miscellaneous | 13 | 5.5 |
| Number of customers | 1-35 | 13 | 13.9 |
|  | 36-70 | 71 | 29.8 |
|  | 71-105 | 88 | 37.0 |
|  | >105 | 46 | 19.3 |
| Average sitting time, h/d | 1-3 | 63 | 26.5 |
|  | 4-6 | 150 | 63.0 |
|  | 7-9 | 25 | 10.5 |
| Average standing time, h/d | 1-3 | 102 | 42.9 |
|  | 4-6 | 126 | 52.9 |
|  | 7-9 | 10 | 4.2 |
| Availability of light | yes | 196 | 82.4 |
|  | no | 42 | 17.6 |

### Logistic regression analysis of study variables

Bivariate analysis of socio demographic characteristics of participants revealed marital status, monthly salary and work experience of participants significantly associated with WMSDs. Laboratory personnels that got a monthly salary of greater than 4500ETB had little risk of experiencing WMSDs than those who earned less (OR, 2.451;95% CI, 1.108-5.420). The more experienced laboratory staffs i.e. > 5 years had lesser experience WMSDs than those who had < 5 years of work experience (OR, 0.063; 95% CI, 0.013-0.2975). Age (OR, 0.147; 95% CI, 0.062-0.347) and Sex (OR, 0.975; 95% CI, 0.578-1.645) did not fit with the model of Hosmer and Lemeshow test. Different Individual characteristics of study participants were analyzed among them reading books as a recreation was greater than two times more likely to develop a risk of WMSDs (OR, 2.451;95% CI, 1.108-5.420). Regular physical exercise had an association with report of WMSDs (OR, 1.399; 95% CI, 0.821-2.381) at a significant value of 0.217. Type of recreation, diagnosis of bone disease had no an association with reports of WMSDs (Table 6).

**Table 6.** Logistic regression analysis factors among MLPs in Bahirdar, Ethiopia, 2021

| Variable | Category | WMSDs |  | COR (95% CI) | P-value | AOR(95% CI) | P value |
| --- | --- | --- | --- | --- | --- | --- | --- |
|  |  | Yes | No |  |  |  |  |
| Marital status | Widowed | 5 | 1 | 1 |  | 1 |  |
|  | Single | 72 | 63 | 4.247 (0.483-37.325) | 0.192 | 1.167(0.594-2.291) | 0.614 |
|  | Married | 38 | 59 | 7.763(0.873-69.047) | 0.066 | 0.141(0.013-1.523) | 0.107 |
| Monthly salary in ETB | <4501 | 32 | 21 | 1 |  | 1 |  |
|  | 4501-7501 | 59 | 62 | 0.873(0.467-1.633) | 0.671 | 1.204(0.566-2.560) | 0.630 |
|  | >7501 | 25 | 33 | 0.497(0.233-1.060) | 0.070 | 1.172(0.424-3.245) | 0.759 |
| Work experience | <5 years | 14 | 2 | 1 |  | 1 |  |
|  | 5-15 years | 76 | 61 | 0.354(0.200-0.626) | 0.000 | 0.325(0.156-0.678) | 0.003 |
|  | >15 years | 26 | 59 | 0.063(0.013-0.297) | 0.000 | 0.056(0.009-0.336) | 0.002 |
| Type of recreation | Home activities | 41 | 29 | 1 |  | 1 |  |
|  | Watching TV | 60 | 67 | 1.579(0.876-2.847) | 0.129 | 1.146(0.574-2.288) | 0.700 |
|  | Reading books | 15 | 26 | 2.451(1.108-5.420) | 0.027 | 2.348(0.957-5.760) | 0.062 |
| Regular physical exercise | No | 70 | 83 | 1 |  | 1 |  |
|  | Yes | 46 | 39 | 1.399(0.821-2.381) | 0.217 | 0.550(0.292-1.037) | 0.065 |
| Repetitive movement within < 30 s | Never | 12 | 19 | 1 |  | 1 |  |
|  | Sometimes | 54 | 66 | 0.467(0.202-1.081) | 0.075 | 0.980(0.397-2.418) | 0.965 |
|  | always | 50 | 37 | 0.772(0.344-1.730) | 0.530 | 1.020(0.385-2.702) | 0.969 |
| Laboratory section | Cl. Chemistry | 10 | 26 | 1 |  | 1 |  |
|  | Haematology | 20 | 21 | 0.256(0.081-0.813) | 0.021 | 0.463(0.157-1.369) | 0.164 |
|  | Bacteriology | 12 | 8 | 0.641(0.129-3.196) | 0.587 | 0.215(0.055-0.834) | 0.026 |
|  | Parasitology | 32 | 23 | 0.445(0.172-1.155) | 0.096 | 0.166(0.057-0.478) | 0.001 |
|  | ART | 4 | 4 | 0.385(0.159-0.930) | 0.034 | 0.689(0.111-4.279) | 0.689 |
|  | Blood bank | 7 | 9 | 0.641(0.184-2.232) | 0.485 | 0.537(0.129-2.243) | 0.394 |
|  | Miscellaneous | 30 | 32 | 0.220(0.088-0.548) | 0.001 | 0.402(0.143 -1.125) | 0.083 |
| # of customers Serviced in a day | 1-35 | 14 | 19 | 1 |  | 1 |  |
|  | 36-70 | 40 | 31 | 0.571(0.248-1.316) | 0.188 | 0.402(0.145-1.118) | 0.081 |
|  | 71-105 | 41 | 47 | 0.845(0.377-1.894) | 0.682 | 0.626(0.232-1.685) | 0.354 |
|  | >105 | 21 | 25 | 0.877(0.356-2.161) | 0 | 0.434(0.144-1.308) | 0.138 |
| Average standing time, h/d | 1-3 | 49 | 53 | 1 |  | 1 |  |
|  | 4-6 | 59 | 67 | 0.231(0.047-1.142) | 0.072 | 1.356(0.638-2.882) | 0.429 |
|  | 7-9 | 8 | 2 | 1.050(0.622-1.771) | 0.855 | 0.346(0.052-2.300) | 0.272 |
| Average sitting time, h/d | 1-3 | 34 | 29 | 1 |  | 1 |  |
|  | 4-6 | 73 | 77 | 1.237(0.686-2.231) | 0.480 | 1.091(0.468-2.540) | 0.840 |
|  | 7-9 | 9 | 16 | 2.084(0.802-5.417) | 0.132 | 2.291(0.609-8.619) | 0.220 |

Ergonomic risk practice like repetitive movement of medical laboratory works within 30 seconds and doing of high workload had significantly associated with WMSDs. On the other hand, there was no association between variables hours for caring children younger than 5 years and report of WMSDs. The association of report of WMSDs and overtime work, regular housework, rest between regular laboratory tasks, and secondary work did not fit with the Hosmer and Lemeshow test model.

The medical laboratory work place also analyzed on the current study. Many laboratory sections had a negative association with reports of the outcome variable. MLPs working in sections of laboratory like Hematology, Parasitology & urinalysis, ART and miscellaneous, and were found lower risk of WMSDs. Whereas there was significant association between number of customers serviced in a day, average standing time, and average sitting time, with WMSDs. There was no a significant association between status of the health facility, and main workstation. The association of report of WMSDs and availability of light did not fit with the Hosmer and Lemeshow test model.

To minimize confounders adjusted odds ratio was necessary. To analyze well, only variables with p<0.25 in the bivariate analysis were calculated in the multiple logistic regression analysis. Work tincture and section of laboratory had a significant association with WMSDs after analyzed using multivariate analysis.

Laboratory personnels whose work experience greater than ranged from 5-15 years (AOR, 0.325; 95% CI, (0.156-0.678) and ≥15 years (AOR, 0.05;95% CI, (0.009-0.336) with significance value of 0.003 and 0.002 respectively, which had less chance of developing WMSDs. Sections of laboratory such as bacteriology and parasitology and urinalysis had AOR of (AOR, 0.215; 95% CI, (0.055-0.834) and (AOR, 0.166; 95% CI, (0.057-0.478) respectively that implies MLPs who served in listed sections had less risk of WMSDs. Marital status, monthly salary, type of recreation, regular physical exercise, repetitive movement within < 30 seconds, number of customers serviced in a day, average sitting and standing time had no significant association with work related musculoskeletal disorders (Table 6).

Abbreviation: ETB Ethiopian Birr, CI confidence interval, COR crude odds ratio, AOR adjusted odds ratio and 1= reference

### Awareness about ergonomics

Seventy-seven (32.4%) participants reported they heard about ergonomics, and the source of information was regular education 30(12.6%) and training 26(10.9%). All who had awareness about ergonomics believe the necessity to applying the principle of ergonomics. Twenty-six (10.9%) respondents reported they exposed to biological and chemical risk majorly. Laboratory managers i.e. 15 out of 27 reported did not heard about ergonomics. The others (12 out of 25) know little about ergonomics, their source of information was from formal education (50%) and training. They reported the importance of ergonomics to grow productivity of the health facility and to keep the professionals health. Nevertheless, eight managers reported there was no interventional measures took place on the work place, four of the 12 declared that there was training about the concept. Ten (37%) managers reported that the duty of applying the principle of ergonomics should be responsibility of MLPs and managers.

### Evaluation of Ergonomic Workstation

To determine the final score, the numbers of ergonomic workstations evaluated were 156. The mean score of less than 2.00 was scored by 54(34.6%) workstations. That clearly indicates poor ergonomic condition. Hence, the overall mean score was 2.28; no workstation got a good status. A mean score greater than 4.00 implies good ergonomic situation (Table 7).

**Table 7.** Mean score evaluation of medical laboratory workstations

| Scores | 1 | 2 | 3 | 4 | 5 | Total Workstations |  |
| --- | --- | --- | --- | --- | --- | --- | --- |
| Workstations | No. (%) | No. (%) | No. (%) | No. (%) | No. (%) | No. (%)<sup>a</sup> | Mean score<sup>b</sup> |
| Standing bench | 7(25.9) | 14(37.0) | 6(22.2) | NA | NA | 27(17.3) | 1.96 |
| Seating bench | 6(22.2) | 12(44.4) | 9(33.3) | NA | NA | 27(17.3) | 2.11 |
| Laboratory chairs | 6(22.2) | 11(40.7) | 10(37) | NA | NA | 27(17.3) | 2.15 |
| Microscopes | 5(18.5) | 15(55.5) | 5(18.5) | 2(7.4) | NA | 27(17.3) | 2.15 |
| Pipettes | 11(40.7) | 12(44.4) | 4(14.8) | NA | NA | 27(17.3) | 1.74 |
| Computer | NA | 4(36.4) | 6(54.5) | 2(18.2) | NA | 11(7.5) | 3.09 |
| Micromanipulation | NA | 4(40) | 4(40) | 2(20) | NA | 10(6.4) | 2.8 |
| Total | 35(23.9) | 68(46.6) | 40(31.5) | 4(2.7) | NA | 156(100) | 2.28 |

NA, not applicable, n=238 <sup>b</sup>poor ergonomic conditions=2.00 or lower, good ergonomic conditions=4.00 or higher. <sup>a</sup>n= 156 respondents worked in health facilities of Bahirdar.

### General workstation

The association of ergonomic hazards and WMSDs may be understated by making a good setup of workplace for workers. ^16^ Observational method was applied to evaluate the workstation. The parameters that we use to analyze the workstations were evenness of workstation, amount of workspace, suitability of design of tools and equipment, working height adjustment, working chair adjustment, the possibility to sit and rest, repetition of sustained work when twisting of the back, neck and hand, availability of light and repetition of similar movements.

The observation showed that 16(59.3%) of working surface area was evenly arranged well. 16(59.3%) of working space was too limited for work movements. The force needed to complete the task depends on significantly on proper design of tools and equipment. The other fact that worsens the working space was 18(66.7%) tools and equipment design was not suitable. The working height of only 14(51.9%) tables was correctly fit with the workers body condition. 19(70.4%) of professionals had a possibility to sit and rest.

Among twenty-seven medical laboratory professionals that participated in the study 24(88.9%) were on sustained work while their back was mildly flexed forward. 22(81.5%) of study participants were perform their repeated work in uncomfortable hand positions. 26(96.3%) study participants were work on sustained work when their arm reaches forward without support. Furthermore, 25(92.6%) study participants work similar work movements repeatedly.

### Standing and Sitting Workstations

A total of 27 standing benches were assessed among them none of them had anti-fatigue matting, all laboratory benches were fixed, the height of 16(59%) laboratory benches fitted with workers, 17(62.9%) benches had ample leg room, 5(18.5%) benches has contact stressor, 25 (92.6%) workstations had no rounded padded edges to reduce contact stress and 24(88.8%) MLPs had necessary materials around their arm’s reach averagely expected within 11 inches. Twenty-seven seating benches were observed. Forty of them (51.9%) had cut outs usable for seated workers.

### Computer workstation

Totally 27 health facility workstations were assessed. 11(40.7%) health facilities has computer workstations among them 7(63.6%) were private health facilities. 8(72.7%) computer workstations had adjustable seats and back support. All computer work stations has ample leg room, only 3(27.2%) seats had foot rest provided, 3(27.2%) computer monitors are out of arm distance, 3(27.2%) computer workstation tables are not at recommended height. 6(22.2%) workstations had a computer monitor at an arm distance. Nineteen (70.4%) workstations had not ample room to accommodate a keyboard and a computer mouse to make employees to rest their side and forearms parallel to the floor. None of them had glare screens on the monitors.

### Microscope workstation

Among medical laboratory professionals who were on microscopy workstation, five out of 27 had rounded chair. 23 (85%) laboratory professionals has excessive neck flexion (>25 degree). Microscopy workstations have no contact stressor between sharp edges and the forearms. 13 (48%) microscopies are pulled out to the end of the workbench to make the work easier, none of the workstations had armrests or padding and 16(59%) workstations had sufficient leg room and professionals rest their feet on laboratory stool as a support.

### Chair workstation

Totally 27 laboratory chairs were assessed; the number of chairs which had adjustable height and backrests was 15(55.6%). A chair with footrest support was 20(70.0%). In addition, chairs that had adjustable armrests that may be removed if they made an obstacle were 21(77.8%).

### Pipette workstation

The other workstation assessed was pipette. The numbers of pipette workstations were 27. Almost all workstations 26(95%) were used routine pipetting by single manual pipette. 14 MLPs were observed on work for more than two hours per day. One health facility uses multichannel pipettes and works more than 4 hours a day. Five (18.5%) laboratory professionals worked in neutral position with a relaxed arm, wrist and shoulder.

### Micromanipulation

Eighty eight percent of the time (7 of 8 times) workstations were performed micromanipulations like decapping and capping tubes easily.

## Discussion

WMSDs and ergonomic risk factors in medical laboratory department of health institutions were source of negative effect on workers’ quality of life, capacity of productivity, pattern of absenteeism, and disabilities on body parts.^17^ One of the least investigated concepts in Ethiopia is the distribution of WMSDs among medical laboratory personals. The result implies that among MLPs, currently WMSDs are the precedence occupational related factor in Ethiopia. The current study tried to assess the magnitude of WMSDs and ergonomic risk practice among MLPs working in Bahirdar health facilities, Ethiopia from February 1, 2021 to June 30, 2021.

The study revealed the magnitude of WMSDs within 12 months was (48.7%) which in line with studies conducted in Addis Abeba (49.4%) among MLPs. ^11^ and study among adults in Ethiopia 35% to 74.5%. ^18^ Rebelliously, the magnitude of the study finding is greater than the study done among MLPs in Nigeria (34.5%). ^17^, MLPs in India (18%). ^19^ and WMSDs in industry workers of Iran (36%). ^20^ The results of the investigation had lower magnitude than two studies in India (66.9%). ^21^ and (100%). ^22^, another study in Saudi Arabia (82%). ^7^ The variation could be because of disparity in demographic difference and the difference in study setting.

The specific body parts have been affected more were lower back 49(20.6%), wrists/hands 39(16.4%) and upper back 33(13.9%). This study was lower than two studies of Saudi Arabia among laboratory workers were lower back (61%), upper back (44%) and also wrists/hands (34%) and the second study with a magnitude of shoulders (33.5%), low back (27.5%), upper back (26.5%), and neck (23.0%) respectively. ^7, 23^ The pain magnitude from India were back pain (53%), neck (39%), wrist (21%), shoulder (21%), heel (14%) & knee (10%). ^24^ Among office workers of Nigerian hospitals were low back (61.1%), neck (43.4%), shoulder (32.1%), upper back (31.5%), hips (30.6%), hand (26.5%), and elbow. ^25^ A possible explanation regarding the disagreement of studies might be due to socio-demographic variation and study setting.

In the current study, specifically lower back pain had a high magnitude of 49(20.6%). Which is comparative with studies in Nigeria healthcare workers^26^, and general surgeons of India. ^27^, a review from Brazil ^16^ and medical practitioners of Saudi Arabia.^28^ A possible explanation of the agreement between the studies may be due to the nature of the procedure that the work needs or the material design used by these health workers. On the other hand, this study finding was lower than a study done in Nigeria among health workers ^29^ and a study conducted among medical laboratory staffs of Sweden determined the prevalence of lower back pain was (25 to 57%). ^30^ The difference may be due to study setting and socio economic variation.

Laboratory personnel whose work experience ranged from 6 to 10 years (AOR, 0.363; 95% CI,(0.181-0.726) and ≥16 years (AOR, 0.083;95%CI, (0.008-0.842) with significance value of 0.004 and 0.035 respectively, had less chance of developing WMSDs when compared with work experience of 1 to 5 years. Younger professionals that had the work experience of 1 and 5 years were more exposed to WMSDs which supported by studies from India and Australia. ^17, 19, 31^ This result is also consistent with study in Saudi Arabia. ^7^ On the contrary, a study from the same country reported that medical practitioners with work tincture of 6-10 years had twice as likely as study participants with 1-5 years of experiences to develop WMSDs. ^28^

Insubordinately, a study done by Agrawal *et al*. reported duration of employment had no association with occurrence of WMSDs. ^19^ On the other side, a study conducted among Iranian health workers reported no association between problems of WMSDs and work experience. ^32^ In other words, WMSDs may meaningfully relate with medical laboratory work experience, lack of training, lack of systematic working habit. In addition, this may be because less experienced personnels may participate in work that is more technical, little experience to handle bad ergonomic conditions and exposed to high workload. As a result, a new direction of outlook could be necessary to solve the occurrence of WMSDs at early age and stages of career.

One of the major factors in WMSDs was sections of medical laboratory departments. This study found an association between some sections of laboratory and WMSDs. Laboratory staffs that were in sections like bacteriology (AOR, 0.218; 95% CI, (0.065-0.727), and parasitology and urinalysis (AOR, 0.384; 95% CI, (0.152-0.974) had lesser risk of WMSDs. The lesser risk may be due to lower workload and in the parasitology and urinalysis section there is more movements to process and reject samples that may decrease poor posture at work, repetitive movements, and presence of working shifts. The association of the above-listed sections of clinical laboratory and WMSDs was generally concur with similar studies in Addis Ababa.^11^ The reason for the agreement might be due to socio-demographic similarities and evaluation tools used.

Based on the current study, awareness about ergonomics had a less likely association with WMSDs. Among 238 MLPs, 77(32.4%) heard about ergonomics. Which is comparable with the study in Nigeria (25.5%).^2^ Even though, there was no updated study on the local context. Ergonomic awareness of this study was greater than two studies conducted in Ethiopia, the first among MLPs 15.4% ^11^ and the second within industry workers ^14^. The difference might be due to variations on study setting and time.

Nevertheless, ergonomics is new to the healthcare ^2^, regular education and training was the major source of an information. But the worse result was half of interviewed, 15(55.6%) laboratory managers reported they had no any idea about ergonomics, which is supported by previous study findings in Ethiopia ^11^. To get the very benefit of ergonomics, laboratory managers had no basic knowledge which leads to poor application.^2^ Likewise, most participants’ awareness of ergonomics at the workplace was unknown. ^3^ The possible explanation may be due to poor in-house training about ergonomics. ^2^ Moreover, weakness of peer teaching and orientation after taking the training. We understand that the best scheme for preventing WMSDs were to strengthen trainings about ergonomics. ^33^

The major relevant issue to the occurrence of MSDs are the working posture of workers, handling of materials, movements performed repeatedly, static work and work related disorders. WMSDs mostly affect neck/shoulder and low back of the body. ^34^ When the work environment was in poor ergonomic design, which leads to awkward postures and work activities that consequently leads to biomechanical stresses on joints, muscles, and tendons. ^33^

In our study, the mean score of the workstations were 2.28 that show poor ergonomic condition and a comparable study of Iran reported a mean score of 2.2. ^35^ It is contrasting with a study from Ethiopia with a mean score of 1.95. ^11^ The deviation from the study of Haile et al. could be due to set up of laboratories and production of ergonomically well-designed materials currently.

Accordingly, the study exposed that many conditions of standing and sitting workstations were poor with a mean score of 1.96 and 2.11 respectively. There was no antifatigue matting in any workstation, 11(40.7%) height of benches did not adjusted, 81.5% of benches had no contact stressor, 13(48.1%) seated benches had cut-out and all benches had fixed height and no supportive shoes used. It is comparable with a study by Haile *et al* in terms of the percentage of rounded edge of contact stressors, easiness of reaching materials on laboratory benches.^11^ Likewise a study conducted in India, reported prolonged standing, the inappropriate height of tables had no association with WMSDs.^19^ The reason for the comparability of the study may be due to the similarity of socio demographic factors.

Moreover, routine activities that performed in medical laboratory departments such as pipetting, microscopy, micromanipulation, and working with biosafety cabinets and non-standardized work place designs exposed MLPs to WMSDs. ^17, 36^ In our study, the number of chairs with lumbar support and adjustability features was eight (72.7%). The number of workstations without foot support was eight (72.7%). The workstations without enough space to hold monitor and mouse was 19(70.4%) that have consequently uncomfortable hand position. All workstations had no antiglares screens, which is in comparison with the study in Ethiopia reported as the laboratory chairs with adjustment feature was 43.8% ^11^, whereas in Indian study chairs with adjustability feature was 51.6%, workstations that had no foot support were 66.7%, laboratory computers with an antiglare screens was 25.0%. ^19^ The difference might be created because of advancements in furnitures technologies in near feature, limited awareness about the importance of the materials, the necessity of the product may be decreased, example antiglare screens because of improvements on buildings.

The magnitude of sufficient legroom in the microscope workplace was 16(59%). In contrast, the study in India exposed 11.1% of participants on microscope workspace had no enough leg and space which directly leads to enhanced exposure of awkward posture. ^19^ Use of microscope at awkward posture for long period of time associated with WMSDs ^37^ and 23(85%) of MLPs has excessive neck flexion that significantly associated with WMSDs. ^28^ On microscopy workstation, there was no contact stressor between sharp edges and the forearm that creates WMSDs of hand and wrists.^38^ The probable explanatory difference may be awareness variation and workplace design difference.

To reduce the occurrence of musculoskeletal disorders having ergonomic requirements of chairs have been very much important. ^19^ In the present study, fifty (55.6%) chairs had adjustable height and backrests, 20(70%) had foot support and 21(77.8%) had removable adjustable armrests, which is contrast with a study from Addis Ababa resulted with (20.0%) had adjustable height and backrests, 12(60.0%) had footrest support and 5(25.0%) had removable adjustable armrests. ^11^ The differences may be due to technological advancement, increment of awareness of MLPs and the buying power of health facilities. When compared with rehabilitation, golden solution to occurrence of WMSDs on workers, employers and society, is applying preventive measures by detecting potentially harmful ergonomic work situations. ^16^

In the present study, 22(81.5%) MLPs working in pipette workstation did not worked in relaxed shoulder and neutral position of arms and wrists. That may cause upper back pain. Which is supported by similar previous study findings in Ethiopia that reported 86.6% of MLPs did not work in neutral position of arms and wrists and with relaxed shoulder. ^11^ This might be due to poor awareness of ergonomics and knowledge of gains of its right among MLPs. ^2^ Moreover, there are limited studies about ergonomic hazards that ultimately lead to musculoskeletal problems. ^3^ The study from India concluded that the percentage of participants with WMSD that were work on pipette in awkward posture was 6.7%. ^19^ The difference might be due to awareness about neutral position of body, the ability to provide ergonomic workstation and work setup.

Above all, to get effective and efficient result, health and safety MLPs at the work environment should be given utmost concern because 80% of decisions related to admission of patients to hospital, prescriptions of medication and discharges need participation of medical laboratories.^39^ Without proper functioning of ergonomics in work place environment and tasks of medical laboratories, the goal of health institution cannot be achieved hence the efficiency of employees decreased.^2^ Managers’ should emphasize good ergonomic design and safe working environment because physical efforts continue to cause serious damage to workers’ health. ^1, 40^

## Strengthening and Limitation of the study

### Strength of the Study

The study covers all laboratory tasks and laboratory professionals. To assure the quality of data, tools of data collection was pretesting in one of tertiary hospital in Amhara region.

## Limitation of the Study

Due to different factors, this study had some limitations and reader should consider them while inferring our finding: it was limited by cross sectional nature of the study design. The same is true for related studies; it might be difficult to distinguish between WMSDs and musculoskeletal disorders of other causes for MLPs.

## Conclusion and Recommendations

The finding of this study confirms, about half of the study participants, one hundred sixteen (48.7%) MLPs suffered from WMSDs and majority was from public hospitals. Among them, 42.4% of MLPs took medication. Of the nine body regions lower back, wrist/hand and upper back pain was the majorly affected. MLPs reported the foremost causes of WMSDs were doing the same laboratory tasks repeatedly within short period of time and awkward posture. The choice of the recreation type for 70.6% of medical laboratory staffs were watching TV and reading books. Doing physical exercise did not a part of majority MLPs lifestyle.

Binary logistic regression analysis revealed marital status, monthly salary, work experience, regular physical exercise, repetitive movement of works within 30 seconds, section of laboratory, number of customers served in a day, average standing time, and average sitting time were significantly associated with WMSDs. Likewise, work experience and sections of laboratory are significantly associated with WMSDs in multiple logistic regression analysis. More broad research into ergonomics and ergonomic risk practice may help to make solutions, consequently to improve the current conditions.

About one third 77(32.4%) of MLPs heard about ergonomics that inversely explained majority of MLPs did not aware about ergonomics at the workplace. More specifically 15(55.5%) laboratory managers did not know heard anything about ergonomics and 12(44.4%) know little about the concept and 37% of managers reported that the duty of applying ergonomics principle should be obligation of MLPs and managers. Training and orientation about causes of WMSDs for especially less experienced MLPs to adopt preventive measures is so urgent.

The workstations scored of standing bench, seating bench, laboratory chairs, Microscopes, pipettes, computer and micromanipulation were noncompliance. The final overall score was 2.28, which showed poor ergonomic workstation. To decrease WMSDs, laboratory workplace setup takes priority. It is essential to make modification of tools and equipments to fit with the body nature of MLPs. We expect our research call the need of action on the matter of causes of WMSDs among MLPs of Ethiopia.

## Data Availability

All data produced in the present work are contained in the manuscript

## Acknowledgments

We thank all health facilities’ and MLPs for participating in the study.

## Authors’ contributions

Mekuriaw Temeche Albsew is the principal author, and contributed to methodology, data collection, data analysis, manuscript preparation and review, appraisal and editing.

Abay Sisay Misganaw contributed to research idea conception, methodology and manuscript review, appraisal and editing.

Melashu Balew Shiferaw contributed to methodology and data collection Habtamu Molla contributed to methodology and data collection

We read and approved the final manuscript.

## Ethical considerations

To do the study, ethical clearance was obtained from DRERC (department research and ethical review committee) of Addis Abeba University, college of health science with a protocol number of DRERC/618/21/MLS. During data collection, oral consent was obtained from study participants by explaining the aim and purpose of the study and their rights.

## Declaration of conflicting interests

The author(s) declared no potential conflicts of interest with respect to the research, authorship, and/or publication of this article

### Data Availability statement

All data produced in the present work are contained in the manuscript

